# Predictors of 90-Day Colectomy in Hospitalized Patients Receiving Infliximab for Acute Severe Ulcerative Colitis at a Tertiary Care Center

**DOI:** 10.1101/2022.05.16.22275155

**Authors:** Jeffrey A. Berinstein, Neelakanta Anurag Atkuri, Elliot M. Berinstein, Jessica L. Sheehan, Laura A. Johnson, Shirley A. Cohen-Mekelburg, Hui Jiang, Nicole I. Wakim, Kelley M. Kidwell, Inbal Billie Nahum-Shani, Robert, Akbar K. Waljee, Peter D.R. Higgins

## Abstract

**BACKGROUND & AIMS:** Acute severe ulcerative colitis (ASUC) is a life-threatening presentation of ulcerative colitis requiring prompt treatment. Infliximab (IFX) rescue therapy can prevent colectomy in patients with ASUC.

**METHODS:** We performed a cohort study of adult patients hospitalized with ASUC between 01/2014 – 02/2021 who received infliximab rescue therapy. Using multivariable logistic regression, baseline and longitudinal laboratory predictors of colectomy within 90 days of index hospitalization were identified and used to develop a risk prediction model and prognostic scoring system to identifying patients at risk for colectomy. Model performance was assessed using area under the receiver operator characteristic curve (AUC) analysis.

**RESULTS:** 166 patients with ASUC who received infliximab rescue therapy were identified. Overall, 24.7% (n=41) underwent colectomy within 90 days. Multivariate analysis revealed that colectomy within 90 days could be predicted by having a calculated IFX clearance of >53L/d (OR 2.28, 95% CI 0.99, 5.22), a day 0 absolute C-reactive protein (CRP) > 91mg/L (OR 3.49, 95% CI 1.58 to 8.08), a decrease in CRP of < 43% from day 0 to day 3 (OR 4.13, 95% CI 1.84 to 9.94), and a decrease in CRP of < 9% from the day of IFX administration to one day post-IFX administration (OR 4.20, 95% CI 1.74 to 10.4). Using these predictors, two highly accurate prognostic scoring system were developed to identify patients at risk for requiring colectomy both prior to administering infliximab (AUC 0.76) and after administer infliximab rescue therapy (AUC 0.81).

**CONCLUSION:** We identified important baseline and longitudinal laboratory predictors of colectomy within 90-days among patients with ASUC receiving infliximab rescue therapy and developed accurate prognostic scores to determine risk of colectomy. These scores can be used risk-stratify patients and facilitate personalized management.

## INTRODUCTION

Ulcerative Colitis (UC) is a chronic gastrointestinal disorder that affects over one million Americans.^1,2^ As many as 30% of patients with UC will require hospitalization for treatment of acute severe ulcerative colitis (ASUC), a life-threatening presentation of UC associated with rapid clinical deterioration, colectomy, and death.^3–5^ First-line therapy consists of rapid induction therapy with intravenous (IV) corticosteroids, however, up to 30% of patients with ASUC will not adequately respond to corticosteroids.^6^ Infliximab and cyclosporine rescue therapies are effective preventing colectomy among patients with ASUC who have failed to respond to intravenous corticosteroids.^7–9^

Despite the use of rescue therapy, ∼30% of patients do not respond to conventional infliximab dosing regimens. Accelerated infliximab induction protocols, where patients receive a second dose of infliximab prior to the typical 14-days, can further reduce the likelihood of colectomy compared to conventional infliximab dosing regimens;^10,11^ However, the optimal dosing and timing of infliximab rescue therapy remains uncertain.^12^ While infliximab levels tend to correlate with treatment response, colonic protein loss in the setting of severely active disease has been associated with lower infliximab levels, likely decreasing infliximab efficacy in patients with ASUC.^13–15^ This is further supported by data demonstrating an association between higher calculated infliximab drug clearance and a higher likelihood of colectomy in patients with ASUC receiving infliximab rescue therapy.^16^ A better understanding of the predictors of infliximab treatment response could help decision making related to ASUC treatment. Current models for predicting treatment response, such as the Oxford Criteria, have largely been developed for corticosteroids alone and do not take into consideration the common use of medical rescue therapies (Ex. the Ho criteria includes cyclosporine rescue, but not infliximab rescue).^17–21^

Therefore, we aimed to accurately identify baseline and longitudinal laboratory predictors associated with colectomy within 90-days of admission for patients with ASUC before and after receiving infliximab rescue.

## MATERIALS AND METHODS

### Study Population

We identified adult patients 18 years or older who were hospitalized with ASUC between January 2014 and February 2021 at a tertiary referral center and received inpatient infliximab rescue therapy. Patients were included if they had an inpatient encounter associated with a diagnosis of UC based on International Classification of Diseases (ICD) −9 and −10 codes and received both intravenous methylprednisolone and infliximab during that hospitalization. Medical charts were reviewed to confirm a diagnosis of ASUC defined by meeting either Truelove and Witts’ criteria, having a C-reactive protein (CRP) ≥ 4.5 mg/dL, or having severe endoscopic disease (Endoscopic Mayo Score ≥ 2).^17,22^ Patients received a single dose of rescue infliximab on day 3 of admission according to a standardized ASUC protocol^23^ which uses well established criteria to identify which patients are high-risk for failing first-line IV corticosteroids.^17,19,20^ Patients who had at least a 20% reduction in CRP three days after their first dose of infliximab and continued to exhibited signs of active disease were offered a second dose if their CRP had not normalized.^23^ The University of Michigan Institutional Review Board (HUM00184362) approved this study.

### Variables and Outcomes

The primary outcome was colectomy within 90 days of the initial hospitalization date. 90-day colectomy was selected to be concordant with prior landmark clinical trials.^7,24^ Patient demographics, UC characteristics, inpatient encounter characteristics, laboratory results, and clinical course and outcome variables were extracted from the electronic medical record (JAB, NAA, and JLS). Infliximab clearance was calculated using a published formula (1) which included albumin concentration (g/dL) at baseline, anti-tumor necrosis factor inhibitor (ATI) status (with 0 signifying the absence of ATI by lab evaluation or at baseline if patient had no prior anti-tumor necrosis factor exposure), and sex (with 1 corresponding to females and 0 to males).^16,25^

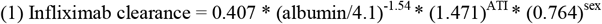

### Statistical Analysis

Descriptive statistics were reported as means and standard deviations for continuous variables with a normal distribution, as medians and interquartile range for non-normally distributed continuous variables, and as frequency and percentages for categorical variables. Missing laboratory values were imputed by random forest imputation method using the {*missforest* package} in R.^26^ Patients who underwent colectomy within the first four days of admission were not included in our analysis, as using a prognostic model to predict an outcome that has already occurred will lead to overfitting (this reduced our sample size by 3). We calculated the area under the curve (AUC) using receiver operating characteristic (ROC) analysis for our baseline and longitudinal laboratory predictors to determine how good each predictor was at classifying between colectomy status. Strong predictors of colectomy (AUC closer to 1) were then evaluated in multivariable regression models. Odds ratios (OR) were used to represent the association between the predictors and outcome of interest and 95% confidence intervals (95% CI) were reported. Models were adjusted for the severity of UC at presentation by the simultaneous inclusion of well-established predictors of colectomy^3,17,27^ and sequential backwards elimination of predictors not significantly associated with our outcome of interest. Multiple models were compared, and the optimal model was selected using the Bayesian information criterion. We also tested whether data collected after before and after INFLIXIMAB dosing would improve the model accuracy. The final model was fit to the entire cohort and model performance (calibration and discrimination) were evaluated using sensitivity, specificity, AUC, and c-statistic. Predictors were evaluated as both continuous and dichotomized variables. Optimal cut-point values for predictor variables were determined according to Youden index using the {cutpointr} package in R which maximizes the sum of the sensitivity and sensitivity of predictor of interest to determine an optimal cut-off value. After fitting the model on the entire dataset, we developed a simplified prognostic scoring system for predicting colectomy risk within 90 days. Our prognostic score was tested with both uniform (1 point per predictor) and variable weighting (based on regression coefficient).^28^ The total risk score was calculated by adding each component, and colectomy rates and colectomy risk (using Cox proportional hazards regression and Kaplan-Meier plots) were determined for the various scores. Probability of colectomy was computed at different cut-points for the total score and three categories of risk (low-risk, medium-risk, and high-risk) were developed. Statistical significance was determined by p<0.05 (two-tailed). All analyses were performed using R version 4.0.3 R Foundation for Statistical Computing, Vienna, Austria).

## RESULTS

### Study Population

Between 2014 to February 2021, 166 patients received intravenous methylprednisolone, followed by rescue infliximab in the hospital for acute severe ulcerative colitis. Baseline characteristics are presented in **Table 1**. Of the 166 included patients, the mean age was 39.5 years, 85 patients (51.2%) were female, 146 patients (88.0%) were white, 130 patients (79.3%) had pancolitis, and 55 patients (33.3%) had reported a prior biologic exposure. Admission characteristics and unadjusted outcomes are presented in **Table 2**. Among the 166 included patients, 16 patients (9.8%) were transferred from an outside hospital, and the mean duration of steroids prior to admission was 15.6 days. The average Truelove and Witts’ Score was 3.96 with 152 patients (91.6%) meeting the Truelove and Witt’s Criteria for ASUC and all 166 (100%) meeting either Truelove and Witts’ criteria, having a CRP ≥ 4.5mg/dL, or having severe endoscopic disease activity. The mean admission C-reactive protein (CRP) was 100mg/dL, the mean baseline hemoglobin was 11.50g/dL (reflecting both anemia and dehydration upon admission), and the mean baseline calculated infliximab clearance was 0.47 L/d. Seventeen patients (10.2%) had colonic dilation, 12 patients (7.3%) had concomitant *Clostridium difficile* and 4 (2.6%) had concomitant cytomegalovirus detected by biopsy. Sixty-four (38.6%) patients received accelerated infliximab with two doses during admission. Over a mean follow-up of 2.30 years, the colectomy rate was 22.3% (n=37) within 30 days, 24.7% (n=41) within 90 days, 29.5% (n=49) within 6 months, and 31.9% (n=53) within 12 months of admission.

**Table 1:** Baseline Characteristics

|  | Overall (N=166) |
| --- | --- |
| Age, years | 39.5 (17.1) |
| Female | 85 (51.2%) |
| Race |  |
| - White | 146 (88.0%) |
| - Black | 9 (5.4%) |
| - Other | 11 (6.6%) |
| Duration IBD, years | 7.0 (10.0) |
| Distribution |  |
| Proctitis | 4 (2.4%) |
| Left-sided Colitis | 30 (18.3%) |
| Pancolitis | 130 (79.3%) |
| Prior Biologic Use | 55 (33.3%) |
| Number of Prior Biologic Exposure |  |
| 1 | 43 (25.9%) |
| 2 | 10 (6.0%) |
| 3 | 6 (3.6%) |
| Prior Infliximab Exposure | 37 (22.3%) |
| Prior Adalimumab Exposure | 25 (15.1%) |
| Prior Vedolizumab Exposure | 13 (7.8%) |
| Prior Ustekinumab Exposure | 0 (0.0%) |
| Prior Tofacitinib Exposure | 4 (2.4%) |

Continuous variables are presented as means (standard deviations) for normally distributed data and medians (interquartile range) for non-normally distributed data. Categorical variables are presented as numbers (percentages).

**Table 2:** Admission Characteristics and Outcomes

|  | Overall (N=166) |
| --- | --- |
| Outside Hospital Transfer | 16 (9.8%) |
| Mean Duration Steroids Prior to Admission (days) | 15.6 (43.1) |
| Truelove Witts Score | 3.96 (1.04) |
| Truelove Witts Met | 152 (91.6%) |
| ASUC Criteria Met | 166 (100.0%) |
| Admission CRP (mg/L) | 100.3 (77.4) |
| Hemoglobin (g/dL) | 11.50 (2.28) |
| Calculated Infliximab Clearance (L/d) | 0.47 (0.15) |
| Colonic Dilation | 16 (9.9%) |
| Endoscopic Mayo Score |  |
| Mayo 2 | 43 (26.7%) |
| Mayo 3 | 118 (73.3%) |
| Unavailable | 5 |
| *Clostridium difficile* | 12 (7.5%) |
| Cytomegalovirus | 4 (2.7%) |
| Accelerated Rescue Infliximab | 64 (38.6%) |
| Initial Infliximab Dose |  |
| 5mg/kg | 52 (31.3%) |
| 10mg/kg | 114 (68.7%) |
| Cumulative Infliximab Dose (mg/kg) | 11.7 (5.3) |
| Length of Stay (days) | 9.8 (7.2) |
| 30 Day Colectomy | 37 (22.3%) |
| 90 Day Colectomy | 41 (24.7%) |
| 6 Month Colectomy | 49 (29.5%) |
| 1 Year Colectomy | 53 (31.9%) |
| Follow-up (years) | 2.3 (1.8) |

Continuous variables are presented as means (standard deviations) for normally distributed data and medians (interquartile range) for non-normally distributed data. Categorical variables are presented as numbers (percentages).

### Univariate Predictors of Colectomy within 90 Days

Data from only 163 patients was used in this analysis, as three patients had colectomy outcomes prior to day 3 of hospitalization. **Figure 1a** illustrates the area of under the curve (AUC) of our baseline and longitudinal laboratory predictors of colectomy within 90 days. Absolute CRP and change in CRP are more accurate predictors of colectomy compared to the calculated infliximab clearance in ASUC patients receiving infliximab rescue therapy. The AUC for the absolute CRP was higher later in the admission with an AUC 0.73 for CRP on day 3. The AUC for the percent change in CRP was highest from day 0 to day 1 with an AUC of 0.61. The AUC was highest when CRP was measured two days after infliximab administration (AUC 0.63).

**Figure 1:**
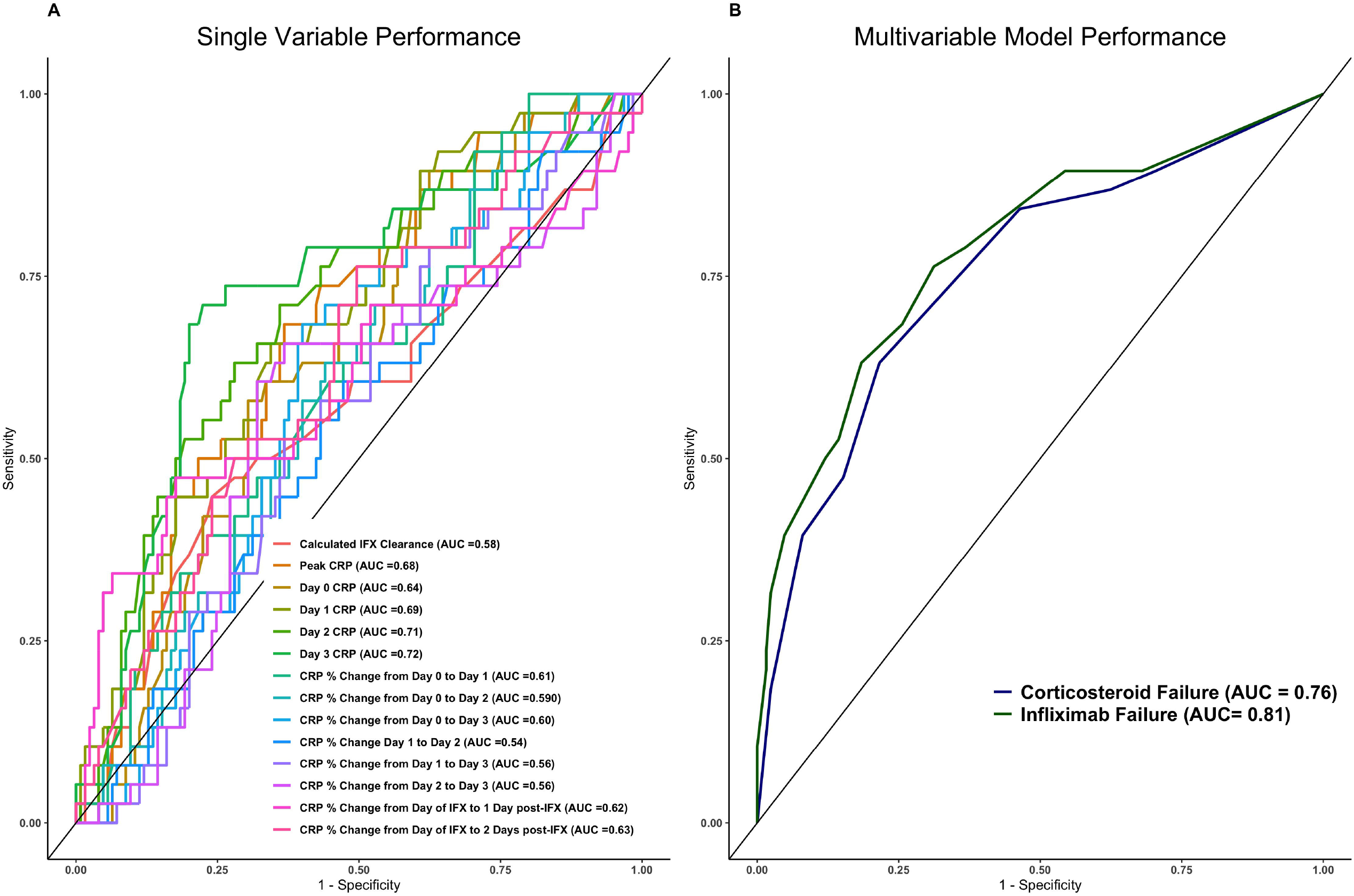
Univariable (1A) and Multivariable Model (1B) Performance at Predicting Colectomy within 90 days of Hospitalizations.

### Multivariable Predictors of Colectomy within 90 Days

Two final models were built to determine risk of colectomy within 90 days both before and after infliximab administration (**Supplemental Table 1**). Predictors were evaluated as both continuous and dichotomized variables. The previously described AUC analysis was used to determine the optimal cut-off values which maximize sensitivity and sensitivity for predicting colectomy within 90 days. In the model predicting corticosteroid failure (prior to rescue infliximab administration), significant predictors included baseline infliximab clearance > 0.53L/day (OR 2.28, 95% CI 0.99 to 5.22, p = .052), a day 0 CRP of > 91mg/L (OR 3.49, 95% CI 1.56 to 8.08, p = .002), and a decrease in CRP < 43% from day 0 to day 3 (OR 4.13, 95% 1.84 to 9.94, p<.001). In the model predicting infliximab failure (after rescue infliximab administration), significant predictors included baseline infliximab clearance > 0.53L/day (OR 2.87, 95% CI 1.18 to 7.13, p = .020), a day 0 CRP of > 91mg/L (OR 3.09, 95% CI 1.32 to 7.43, p = .009), a decrease in CRP < 43% from day 0 to day 3 (OR 3.66, 95% 1.57 to 9.13, p=.002), and a decrease in CRP <9% from the day of infliximab administration to 1-day post-INFLIXIMAB administration (OR 4.20, 95% CI 1.74 to 10.4, p=.001). The AUC of our corticosteroid failure model was 0.76 and the AUC of our INFLIXIMAB failure model was 0.81 (**Figure 1B**).

### Simplified Corticosteroid Failure Prognostic Score

Our corticosteroid failure model included: baseline infliximab clearance > 0.53L/day, a day 0 CRP of > 91mg/L, and a decrease in CRP < 43% from day 0 to day 3. The same weight for all variables was chosen with 1 point for each item. This performed better than a slightly more complex model with weights corresponding to coefficients. In our corticosteroid failure model, a score of 0 was considered low-risk, and a score of 1 was considered moderate-risk and a score 2 or 3 was consider high-risk. Proportions of patients undergoing colectomy and rate of colectomy-free survival for each risk score category are illustrated in **Figure 2A** and **Figure 2B**, respectively. 4 (10%) patients in the low-risk group (n=41), 10 (14%) in the moderate-risk group (n=71) and 24 (47%) in the high-risk group (n=51) underwent colectomy with an associated hazard ratio (HR) of 1.50 (95% CI 0.47 to 4.79, p = .50) and HR of 6.20 (95% CI 2.15 to 17.9, p < .001) for the moderate-risk and high-risk group, respectively.

**Figure 2:**
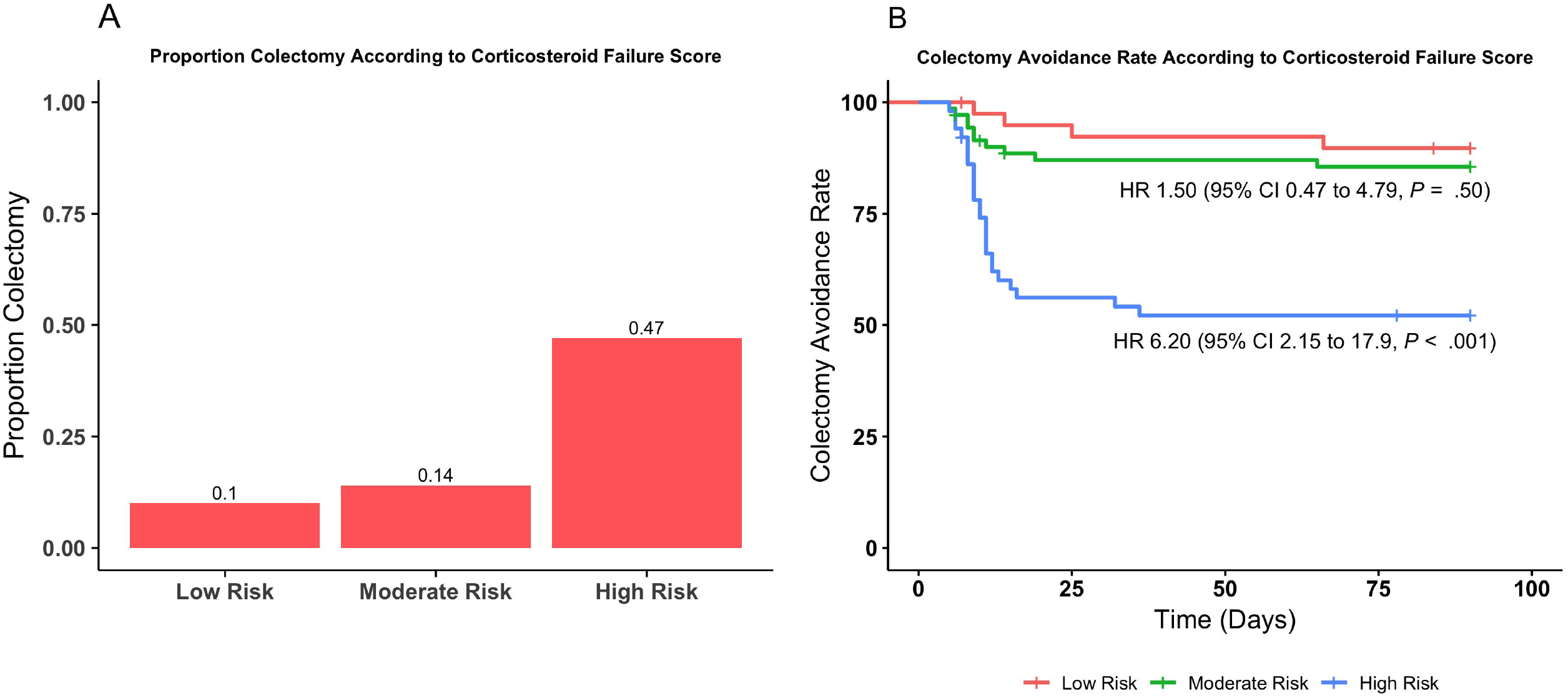
Corticosteroid Failure Prognostic Score Performance According to Proportion Colectomy (2A) and Calculated Hazard Ratio (2B).

### Simplified Infliximab Failure Prognostic Score

Our infliximab failure model included baseline infliximab clearance > 0.53L/day, a day 0 CRP of > 91mg/L, a decrease in CRP < 43% from day 0 to day 3 as well as a CRP < 9% from the day of infliximab administration to 1-day post-infliximab administration. The same weight for all variables was chosen with 1 point for each item. This performed better than a slightly more complex model with weights corresponding to coefficients. In our infliximab failure model, a score of 0 and 1 was considered low-risk, and a score of 2 was considered moderate-risk and a score 3 or 4 was consider high-risk. Proportions of patients undergoing colectomy and rate of colectomy-free survival for each risk score category are illustrated in **Figure 3A** and **Figure 3B**, respectively. 9 (10%) patients in the low-risk group (n=95), 14 (30%) in the moderate-risk group (n=47) and 15 (71%) in the high-risk group (n=21) underwent colectomy with an associated HR of 3.62 (95% CI 1.56 to 8.36, p = .003) and HR of 12.4 (95% CI 5.40 to 28.7, p < .001) for the moderate-risk and high-risk group, respectively.

**Figure 3:**
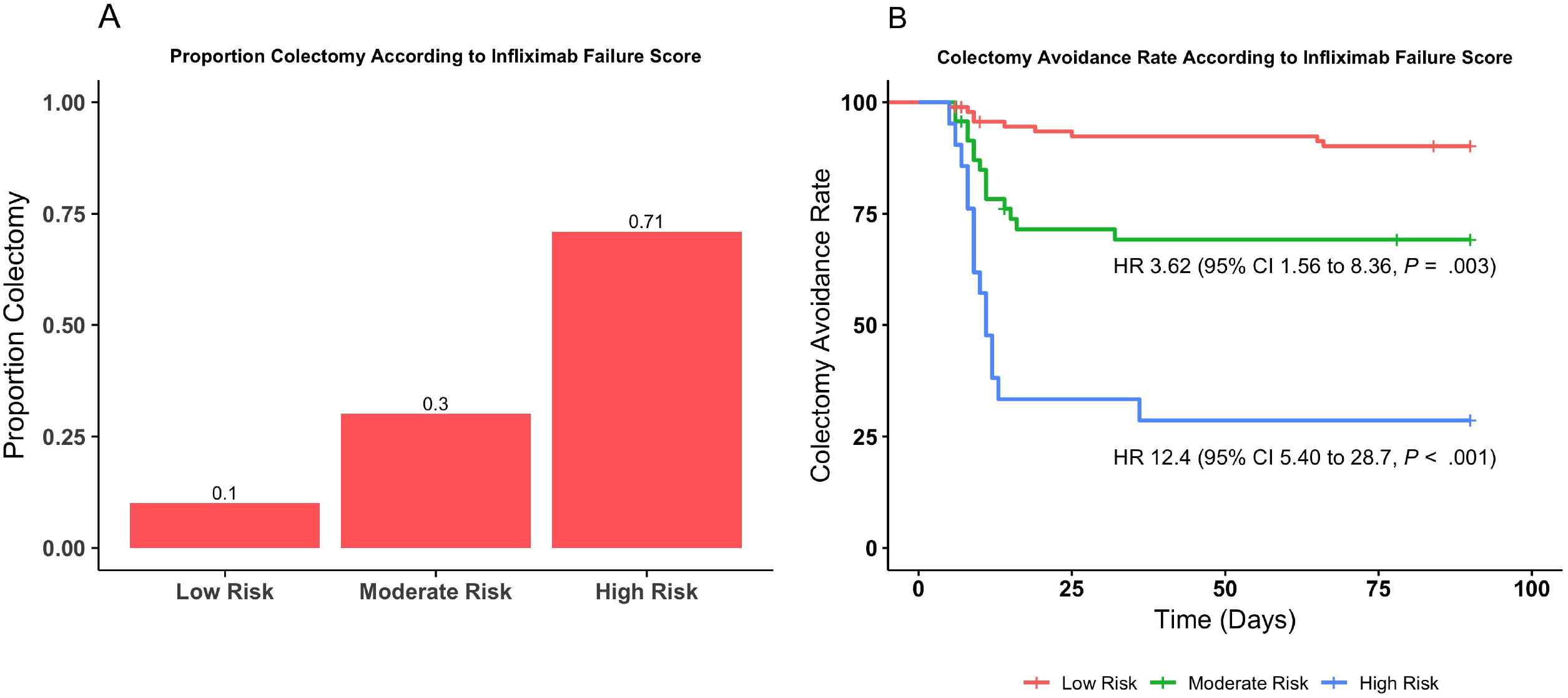
Corticosteroid Failure Prognostic Score Performance According to Proportion Colectomy (3A) and Calculated Hazard Ratio (3B).

## DISCUSSION

In this study we present our experience using infliximab rescue for acute severe ulcerative at a large tertiary care center. Among 166 patients admitted to our institution with ASUC, the colectomy rate was 24.7% (n=41) within 90 days of hospitalization. We identified and evaluated multiple baseline and longitudinal laboratory predictors of colectomy including infliximab clearance, as well as absolute CRP and change in CRP at various timepoints throughout the index hospitalization. Using this data, we were able to build two parsimonious multivariable model which informed the development of two prognostic scores identifying patients at risk for colectomy within 90 days both before and after infliximab administration. Based on our data, baseline infliximab clearance > 0.53L/day, a day 0 CRP of > 91mg/L, a decrease in CRP < 43% from day 0 to day 3 as well as a CRP < 9% from the day of infliximab administration to 1-day post-infliximab administration were the best predictors of colectomy within 90 days. Our models were highly accurate for predicting colectomy with an AUC of 0.76 for the corticosteroid failure model (prior to infliximab administration) and 0.81 for the infliximab failure model (after infliximab administration). Of note, budling multivariable models significantly enhanced prognostic performance (AUC 0.76-0.81) compared to each univariable predictor (max AUC 0.72) alone.

Our study is unique for several reasons. First, the majority of published prediction scoring systems currently in use were developed in the context of corticosteroid or cyclosporine use.^17,19,29^ Even among modern prediction scores, including the one recently derived by the Saint Antoine IBD Network consisting of 270 patients admitted to two hospitals in France with ASUC. only 10 patients (3.7%) were treated with infliximab rescue therapy. Therefore, it is unknown whether these current risk scores provide an accurate estimate of the risk of colectomy in the context of modern rescue therapies.^18,29^ Second, our study looked at a combination of baseline and longitudinal laboratory predictors of treatment response. Accounting for the evolution of patient risk for colectomy over the course of the admission more accurately represents the temporal dynamics of relevant risk factors to more closely mimic real-world healthcare settings. In addition, this will allow us to determine the optimal timing and cut-off of predictors which could be used to identify non-responders early more accurately and administer rescue pharmacotherapy or send for salvage colectomy without missing their “treatment window”. Third, our models are unique in that they calculate risk for colectomy both before and after rescue infliximab administration, as the majority of the models utilized in practice predict colectomy without consideration response to rescue therapy. Fourth, we provide real-world outcomes on the effects of infliximab administration in a large cohort of patients who received rescue infliximab.^30^ Our cohort is representative of the patients typically cared for at tertiary care centers in the United States.^29,31^

In our study, four criteria were associated with colectomy within 90 days in multivariable analysis. First, calculated infliximab clearance based on sex, albumin, and the presence of anti-necrosis factor inhibitors has been shown in multiple studies to be associated with the need for colectomy in patients with ASUC.^16^ In a study by Battat *et al*., a cut-off of 0.627 L/d was used, however in our study a cut-off of 0.53L/d was found to better classify colectomy status in our cohort. The observed difference in cut-off is likely related to differences in study populations. Our second, third, and fourth criteria, involve longitudinal measures of CRP namely an absolute cut-off on day 0 of > 91mg/L, a decrease in CRP of < 43% from day 0 to day 3 as well as a CRP < 9% from the day of infliximab administration to 1-day post-infliximab administration. CRP is a widely used as tailoring variable to base decisions on rescue medication administration and referral to surgery. The Oxford criteria, published in 1996, showed that having greater than eight stools per day or three to eight stools per day and a CRP >45 mg/L on day 3 was associated with an 85% chance of requiring colectomy.^17,19^ However our models unique in that it considers both an absolute CRP cutoff on day of admission and percent change in CRP from day 0 to day 3 and that it looks at response to corticosteroids alone and corticosteroids plus rescue infliximab. This allows us to account for patients with a higher starting value and those with adequate improvement without committing them to colectomy if they are on the right trajectory but not falling below an absolute cut-off. Better incorporation of time-varying factors using longitudinal risk stratification machine learning techniques could be used to develop an early warning system to identify patients at risk of deterioration and offer early and a personalized intervention. Classifying patient response profiles according to the risk of failing first-line and rescue treatment could allow us to personalize our approach by initiating therapy with a different mechanism of action. This overall approach would allow providers to tailor the timing and mechanism of therapy to a patient’s individual health needs.^32,33^

Our corticosteroid failure and infliximab failure prognostic scores developed in this study has several strengths. First, they are easy to use since it only contains four easily and routinely measured dichotomous variables. Second, the scores were developed in a relatively large cohort with a high AUC. Third, our study identifies important predictors of response to medical therapy in a largely understudied population, patients receiving infliximab rescue, despite widespread use of infliximab rescue in clinical practice.

Our study has several weaknesses. First, our study findings may not be generalizable to other centers since the data are derived from a single large tertiary care center. Second, potential residual confounding due to patient and physician preference cannot be excluded. While we developed a public ASUC protocol in an attempt to standardize care at our center, there were deviations from the protocol based on patient or provider preference. Third, model accuracy might have been improved if we were able to reliably incorporate clinical parameters (such a bowel movements, abdominal pain, etc.); however, these variables are often poorly recorded in the electronic health record and not amenable to automated inclusion into our scoring system. Better incorporation of patient reported outcomes throughout inpatient stays could be valuable for future studies in this area.^34^

In conclusion, in this cohort of 163 analyzed patients received infliximab rescue for acute severe ulcerative colitis at a single tertiary care center, several important baseline, longitudinal laboratory predictors associated with colectomy within 90 days were identified. Based on predictive modeling, four variables including calculated infliximab clearance of > 0.53L/day, a day 0 CRP of > 91mg/L, a decrease in CRP < 43% from day 0 to day 3 as well as a CRP < 9% from the day of infliximab administration to 1-day post-infliximab were incorporated into two accurate and easy to use prognostic scoring system that was highly predictive of the colectomy within 90 days of admission. Additional prospective studies are required to confirm the generalizability of these findings.

## Data Availability

All data produced in the present study are available upon reasonable request to the authors

## Abbreviations

ASUC: Acute severe ulcerative colitis
ATI: anti-tumor necrosis factor inhibitor
CI: Confidence intervals
CRP: C-reactive protein
EHR: Electronic medical record
IV: Intravenous
HR: Hazard ratios
UC: ulcerative colitis

