## Supplemental Table 1 for "Predictors of 90-Day Colectomy in Hospitalized Patients Receiving Infliximab for Acute Severe Ulcerative Colitis at a Tertiary Care Center"

### Supplemental Table 1: Multivariable Models

|  | **Corticosteroid Failure Model**  (before IFX administration) | | | | | **Infliximab Failure Model**  (after IFX administration) | | |
| --- | --- | --- | --- | --- | --- | --- | --- | --- |
|  | OR | | 95% CI | p-value | OR | | 95% CI^1^ | p-value |
| **Infliximab clearance >53 L/d** | | 2.28 | 0.99, 5.22 | 0.052 | 2.87 | | 1.18, 7.13 | **0.020** |
| **Day 0 CRP >91mg/L** | | 3.49 | 1.56, 8.08 | **0.002** | 3.09 | | 1.32, 7.43 | **0.009** |
| **Decrease in CRP < 43% from day 0 to day 3** | | 4.13 | 1.84, 9.94 | **<0.001** | 3.66 | | 1.57, 9.13 | **0.002** |
| **Decrease in CRP < 9% from day of IFX to 1 day post-IFX** | | - | - | - | 4.20 | | 1.74, 10.4 | **0.001** |

CI = Confidence Interval; HR = Hazard Ratio; CRP = C-reactive protein; IFX = Infliximab
